# Diagnostic accuracy of fractional exhaled nitric oxide (FeNO) with or without blood eosinophils in childhood, adolescent, and adult asthma: protocol for a systematic review and meta-analysis

**DOI:** 10.1101/2024.07.14.24310394

**Authors:** Marc-André Roy, Morgane Gronnier, Manisha Ramphul, Pip Divall, Samuel Lemaire-Paquette, Andréanne Côté, Francine M Ducharme, Erol A Gaillard, Simon Couillard

**Author notes:** **Address for correspondence:** Dr Simon Couillard, Département de pneumologie et de médecine respiratoire du CHUS, Université de Sherbrooke, 3001 12e Avenue Nord, Sherbrooke (Québec), QC J1H 5N4, Canada.

## Abstract

**Introduction:** Asthma poses a diagnostic challenge due to its intermittent symptoms and variable airflow obstruction. Diagnostic assessments such as spirometry and bronchodilator response are frequently non-diagnostic, necessitating confirmatory bronchial provocation testing. Biomarkers of type-2 inflammation —exhaled nitric oxide (FeNO) and blood eosinophil counts (BEC)— are useful in asthma, but their diagnostic values in children and in combination (FeNO+BEC) are unclear. This systematic review will evaluate the diagnostic accuracy of FeNO alone or in combination with BEC in paediatric and adult asthma.

**Methods and Analysis:** This protocol is reported in line with the Preferred Reporting Items for Systematic Reviews and Meta-Analysis Protocols. The review will include studies of any design on the diagnostic accuracy of FeNO with or without BEC in asthma compared to the reference standards bronchodilator response and provocation testing in patients ≥ 5 years old selected from MEDLINE, Embase, and Cochrane CENTRAL databases. Screening, study selection, and data extraction will be independently performed by two reviewers. Risk of bias will be assessed using QUADAS-2 and QUADAS-C. Meta-analysis will be carried out by pooling the sensitivity and specificity of FeNO alone or in combination with BEC in a bivariate random effects model allowing the generation of summarised operating characteristic curves and summary points. Further analysis utilising a multiple thresholds model will enable the computation of diagnostic thresholds for FeNO.

**Ethics and Dissemination:** No patient data will be stored without prior approval from ethics committee. The findings will be submitted in a peer-reviewed publication.

**Registration:** PROSPERO CRD42023489738

**SOCIAL MEDIA MESSAGE:** Diagnosing asthma is challenging. Spirometry and bronchodilator reversibility are often non-diagnostic, calling for provocation testing. Our systematic review will explore complementary approaches using FeNO with and without the blood eosinophil count.

## INTRODUCTION

Asthma is characterised by variable airflow obstruction, bronchial hyper-reactivity, and chronic airway inflammation, often presenting with variable and non-specific symptoms.[1] Despite being the most common chronic disease in childhood, asthma presents a significant diagnostic challenge. Achieving accurate diagnosis is crucial for effective disease management; however, misdiagnosis is widespread in asthma, resulting in up to 30% of adults and 30-35% of children being inappropriately treated for a condition they do not have[2–4].

The diagnostic standards rely on pre-bronchodilator (BD) spirometry and bronchodilator response (BDR) as initial investigations [5–7]. However, these tests are not diagnostic for many patients who have values within normal references at the time of testing. Indeed, in individuals with a self-reported physician diagnosis of asthma, absence of bronchodilator reversibility had a negative predictive value of only 57% to exclude asthma [8,9]. Bronchial provocation tests (BPT) serve as a highly sensitive secondary measure to confirm diagnosis, but their often-limited availability, significant costs, and procedural risks render them suboptimal as a diagnostic tool for all asthma patients[8].

Recognising these limitations, alternative biomarkers of asthma have emerged as potential screening tools, notably fractional exhaled nitric oxide (FeNO)[5,6,10] alone or in combination with blood eosinophil count (BEC)[11]. These biomarkers reflect the underlying processes of the type-2 inflammatory phenotype of asthma: NO is released by the interleukin (IL)-13-stimulated airway epithelial cells, while IL-5 stimulated circulating eosinophils are an important cell mediator in the inflammatory cascade[12]. Furthermore, these biomarkers can be readily accessible in primary care; FeNO through point-of-care handheld devices [13], and BEC via any general laboratory offering blood cell count from venous blood or capillary samples.

Evidence increasingly supports FeNO as a complementary non-invasive tool for asthma diagnosis. European[5–7] and American societies[13,14] propose FeNO >50 ppb as a diagnostic threshold with specificity >90% for asthma in adults[6], while in children, FeNO >25 ppb is strongly indicative of asthma[5,7], though this threshold remains insufficient for a definitive diagnosis due to its lack of specificity (0.81). Diagnosing asthma solely through fractional exhaled nitric oxide (FeNO) levels is not common practice, as only a minority of adult asthma cases meet its diagnostic threshold, and no established threshold exists for children in current guidelines. Consequently, elevated FeNO levels are primarily employed to bolster clinical suspicion of asthma, without reducing the necessity for further testing in children[5]. In contrast, BEC lacks specificity and sensitivity as an independent diagnostic biomarker and is not recommended for diagnosis alone at any threshold. [5,6,15]. Despite BEC’s limited effectiveness as a standalone diagnostic tool, its combined assessment with FeNO may improve diagnostic accuracy, with additional prognostic (predicting asthma attacks and lung function and theragnostic utilities (predicting response to anti-inflammatory medications)[16,17].

Whereas the existing literature predominantly evaluates the diagnostic accuracy of FeNO [15,18–21] and BEC separately in adult and paediatric populations, there has yet to be a comprehensive evaluation of these biomarkers’ combined diagnostic accuracy for asthma. Nonetheless, emerging evidence suggests that combining FeNO with BEC may enhance its diagnostic accuracy. For instance, in a retrospective cohort study by Li et al. (2021) [22] on adults ≥ 18 years of age with suspected asthma, FeNO levels > 40 ppb combined with BEC > 300 cells/μl, supported an asthma diagnosis with a specificity ≥ 95% and a positive likelihood ≥ 10 with an estimated disease prevalence of 35% when compared to methacholine provocation or BDR. The objective of this systematic review and meta-analysis is to fill this gap in the literature by comprehensively evaluating the combined diagnostic accuracy of FeNO alone or in combination with BEC for asthma with subgroup analyses in children, adolescents, and adults.

In addition, we aim to analyse the diagnostic accuracy of FeNO alone for both adults and children using a multiple thresholds model. While Schneider et al. (2017) [21] have conducted this analysis for adults, we seek to extend this work to include children for the first time. We plan to update the previous multiple thresholds model analysis for adults as a significant body of literature on FeNO has been published since [16, 22]. Our objective is to better define diagnostic thresholds for FeNO in adults and establish a diagnostic threshold for children with the multiple thresholds model.

### Objectives

The primary goal of this systematic review is to review the evidence concerning the diagnostic performance of FeNO alone or in combination with BEC in individuals aged ≥ 5 years with suspected asthma.

The secondary aims included identifying FeNO thresholds using a multiple thresholds model, both alone and in combination with BEC, for different age groups (children aged 5 to ≤11, adolescents aged 12 to ≤17, and adults aged 18 and above). These thresholds would aim to reliably confirm an asthma diagnosis without the need for bronchial provocation testing in patients with spirometry within normal limits.

## METHODS

This protocol has been prospectively registered with the International Prospective Register of Systematic Reviews (PROSPERO; <u>CRD42023489738</u>) and is reported in line with the Preferred Reporting Items for Systematic Reviews and Meta-Analysis-Protocols (PRISMA-P) guidelines[23]. Amendments made to the protocol will be recorded and included in dissemination.

### Inclusion criteria

A. *Design: randomised controlled trials,* cross sectional, cohorts, case-control and case series studies will be considered for inclusion.
B. *Participants:* patients ≥ 5 years old presenting with asthma-like symptoms with previous spirometry results within normal limits (i.e., FEV_1_ or FEV_1_/FVC ratio Z-score > −1.64).
C. *Index test:* included studies must assess baseline FeNO using any device, in accordance with established standard procedures, that is, respecting an acceptable expiratory flow rate (i.e. 50ml/s) and exhalation time (i.e. ≥ 10 seconds, although ≥ 6 seconds is acceptable in children)[24] or baseline BEC on manual or automatic peripheral blood count. Studies must have data detailing specificity and sensitivity of FeNO alone or combined with BEC at defined thresholds, allowing for the construction of 2 × 2 matrix tables for asthma diagnosis compared to the reference standard.
D. *Allowable co-testing:* studies will be considered for inclusion independent of co-testing
E. *Reference standard:* To ensure the identification of studies with robust reference standards, enabling the tabulation of highly specific thresholds, our senior team members will evaluate the diagnostic pathways of eligible studies. The reference standard should include a bronchial provocation test, either direct or indirect, to definitively confirm or infirm asthma in participants. Consequently, the primary gold standard diagnosis will consist of a direct methacholine provocation test with a PC_20_ ≤8mg/ml or PD_20_ ≤200 mcg, or other direct and indirect provocation tests in accordance with European (preferred) and/or American guidelines (complementary)[25]. The eligibility of direct methacholine tests with higher thresholds such as PC_20_ ≤16mg/ml and PD_20_ ≤400 mcg will also be considered in further sensitivity analyses [26]. Spirometry with a bronchodilator response (BDR) is considered adequate for confirming an asthma diagnosis if it shows a variation of ≥12% and, specifically for adults, an increase in FEV_1_ of ≥200 ml from baseline in pre/post-BD testing (“standard criteria” for secondary analysis). Alternatively, a variation of >10% compared to the predicted values for FEV_1_ or FVC in pre/post-bronchodilator spirometry from baseline is also acceptable (“alternative criteria” for secondary analysis). *Although BDR will be included as a reference standard, it will be considered as a secondary exploratory outcome due to its lower sensitivity in asthma.

### Exclusion criteria

A. Studies in which participants have used oral corticosteroids (OCS) in the preceding 2 weeks[27] or have had an upper respiratory virus in the preceding week will be excluded unless subgroup data pertaining to individuals without OCS or URTI are available.
B. Studies in which participants have been identified as current smokers (having smoked in the past 48 hours) [28], at the time of FeNO or BEC testing, will be included in the primary analysis but excluded in a sensitivity analysis [29].
C. Editorials, commentaries, case reports, conference abstracts, reviews, animal, and in vitro studies will be specifically excluded.

### Data sources

A comprehensive search strategy will encompass the following databases: Embase, MEDLINE, Cochrane CENTRAL, ISRCTN registry, UK Clinical Trials Gateway, and Clinicaltrials.gov. Each database will be explored from its inception and restricted to only English and French language publications. The reference lists of all included studies as well as current American[14], European[5,6], Australian [30], New Zealand[31], and International[1] asthma guidelines will be reviewed. Our focus will be on published studies and update searches will be conducted within six months of publication. The draft search strategy (Supplementary material S1) was developed in collaboration with a specialist librarian (P.D.). The search strategy encompassed studies published in English from the inception of the databases until June 2024. See supplementary materials for draft search strategy. MeSH terms and related keywords like FeNO, eosinophils, asthma, diagnosis, and accuracy were employed.

### Study records

Articles from electronic databases and manual reference searching will be compiled in the citation management tool *Covidence*. It will handle citations, references, and bibliographies and enables the identification and removal of duplicates, along with the storage of abstracts and full texts.

#### Selection and Data collection process

Titles and abstracts will undergo screening to identify potentially suitable studies. Subsequently, full texts of articles meeting initial criteria will be acquired and assessed against predefined inclusion and exclusion criteria. Data extraction will be executed using standardized collection forms, initially tested on a subset of eligible articles. Two review authors will independently conduct title and abstract screening, full-text evaluation, and data extraction. Discrepancies will be resolved through consensus among the reviewers. In cases where consensus isn’t reached, a third author will be consulted.

### Data items

We will extract the following data where reported:

A. Study characteristics: including title, year and journal of publication, country of origin, and sources of funding; the first author will be used as the study identification.
B. Participants’ characteristics: age distribution, median age, height, BMI, and assigned sex at birth as well as current inhaled corticosteroids (ICS) / Leukotriene receptor antagonists (LTRA) / long-acting anti-muscarinic agent (LAMA) / oral corticosteroid (OCS) therapy, smoker status, nasal polyposis, atopic comorbidities (atopic dermatitis, allergic rhinitis, allergic conjunctivitis, drug and food allergies and aeroallergen sensitization on prick test or specific serum IgE) and other documented confounding factors as per ERS Monograph : Exhaled Biomarkers (2010) [24].
C. Study Settings: Collection of contextual details about healthcare settings, including distinctions between primary and secondary care.
D. *Index Tests:* Identification and extraction of reported cut-offs for FeNO alone or combined with BEC including their associated sensitivity, specificity, and overall accuracy as well as the counts for true positives (TP), false positives (FP), true negatives (TN), false negatives (FN) for each cut-off and the documented FeNO measuring device and method to count blood cells.
E. *Reference standard details:* Documentation of the diagnostic methodology employed to confirm asthma diagnosis (e.g. such as spirometry, BDR, BPT, or other established means).

### Outcomes and prioritisation

Primary outcome: asthma diagnosis per direct or indirect provocation testing using standard approved criteria, dichotomized as positive or negative.

Secondary outcomes: asthma diagnosis per BDR standard criteria (≥12% change in FEV_1_ and, if adults >200 mL change in baseline FEV_1_) or alternative criteria (>10% change in baseline FEV_1_ compared to predicted FEV_1_), dichotomized as positive or negative, will be investigated as an exploratory outcome.

### Assessment of methodological quality

Risk of bias within the included studies will be independently evaluated by two reviewers utilizing QUADAS-2[32] and QUADAS-C[33] for comparative accuracy studies. These tools encompass four domains – patient selection, index test, reference standard, and flow and timing – each assessed for bias as ‘high’, ‘low’, or ‘unclear’, contributing to an overall judgment of ‘at risk’ or ‘low risk’ for each study. For the flow and timing domain, studies will only be rated as low risk if they have ≤ 6 months between index test and reference standard. Any discrepancies in assessments will be resolved through discussion and consensus between the two reviewers, with involvement from a third author if necessary.

### Statistical analysis and data synthesis

FeNO is a continuous biomarker, and consequently, many studies report accuracy at multiple diagnostic thresholds. Given that each study presents sensitivity and specificity at varying thresholds for the index tests, conducting analyses of data across all thresholds would yield a more clinically informative estimate of their diagnostic accuracy. Therefore, we will synthesise data using the model of Steinhauser et al.[34] where possible.

The Steinhauser multiple thresholds model [34] establishes correlations between varying thresholds and their corresponding sensitivity and specificity pairs, enabling the identification of optimal test thresholds. At the study level, specificities for disease-free individuals and sensitivities for patients with confirmed asthma at available thresholds will be used to estimate the cumulative distribution function (CDF) of non-disease and disease populations based on FeNO. At the meta-analytic level, a log-logistic model (DIDS* which stands for different random intercepts and different random slopes) will be used to estimate the population distribution functions. This will allow the generation of a multiple thresholds summary ROC (mtsROC) curve. For the Steinhausser model, the R programming package: *diagmeta* [35] will be used.

The Steinhauser multiple thresholds model [34] cannot compute threshold based on the combination of FeNO and BEC as it considers only one variable. Consequently, for the combination of both index tests, we will synthesise data using a bivariate random effects model of sensitivity and specificity. Since we expect variations in reported thresholds, we will use the equivalence of the bivariate and hierarchical summary ROC (HSROC) models to generate summary ROC (SROC) curves and summary points for different thresholds or forks of thresholds (e.g., 10-19 ppb, 20-29 ppb, etc) [36]. This approach allows estimation of summary points for particular thresholds of interest, preserving the possibility of comparing the estimated sensitivity and specificity of different thresholds. For the HSROC model, the R programming package: *mada* [37] will be used. We recognise this approach relies on having a substantial amount of data for each threshold, as such this portion of the analysis may be dropped for certain subgroups if data is insufficient in favour of an SROC model using a single threshold per study[38].

The clinical meaning of the index tests diagnostic accuracy will be illustrated using Bayes’ theorem. Disease prevalence will be calculated for each study and across all studies, enabling the calculation of positive and negative predictive values at different thresholds. This process will allow the selection of threshold values for FeNO alone or combined with BEC which have a specificity > 90%, a likelihood ratio > 10, and/or a positive predictive value > 80%.

### Investigations of heterogeneity and sensitivity analyses

Heterogeneity arising from confounding factors will be assessed through subgroup and sensitivity analyses. Subgroup analyses will involve examining studies with vs. without (i) a significant percentage (>25%) of patients with atopic comorbidities (e.g., allergic rhinitis and eczema) or nasal polyposis [13], (ii) inclusion of participants identified as current smokers or utilizing ICS/LTRA therapy[24], and (iii) different age groups (children, adults, or mixed) [25]. Sensitivity analysis will be done on the study design (e.g. retrospective vs. prospective studies) and alternative cut-off values for provocation testing (i.e., positive methacholine at PC_20_ >8mg/ml or PD_20_ >200 mcg). We will consider an I^2^ value of 50% or higher as an indicator of significant heterogeneity[39].

Dealing with missing data is crucial. If required, our team will contact authors or sponsors of included studies to clarify the methodology and/or to obtain missing data on participants or threshold values that have not been reported in their articles. Should individual patient data be required, protocol amendments and ethics committee approval would be necessary before going forward.

### Meta-bias(es)

We acknowledge the potential for biases inherent in the selective reporting of studies and publication bias within the field. We will report the funnel plot for the main outcome and perform Egger’s regression [40] test to evaluate for publication bias.

To provide transparency and address potential biases in study selection, we will present a table outlining the primary characteristics of studies included in our analysis and of those that were deemed potentially eligible but were subsequently excluded, along with the criteria justifying their exclusion. [13]

## DISCUSSION

The landscape of asthma is currently in a transformative phase, marked by significant advancements in our understanding of immune pathways and the emergence of innovative immunological treatments. However, there is a noticeable lag in the development of diagnostic methods. Given the limitations in accessibility and screening associated with traditional diagnostic techniques [2,3,8], our efforts to review alternative diagnostic methods are crucial.

In keeping with existing analyses, our work represents an important update in the field of asthma diagnosis, reflecting the substantial body of literature and data published in recent years [15,22]. Furthermore, local studies – *e.g.* DIVE (clinicaltrials.gov <u>NCT05992519</u>) and DIVE2 (registration pending) – are ongoing prospective cohort studies exploring FeNO and BEC’s combined diagnostic accuracy in adults and children, respectively. Our analysis serves as a foundational reference for these studies which present opportunities to incorporate emerging data into our analysis.

The strengths of this systematic review include its comprehensive examination of the diagnostic accuracy of FeNO, both with and without BEC, across various age groups, and its use of a multiple thresholds model to better synthesize data on FeNO’s diagnostic accuracy, which is often reported at different thresholds. To our knowledge, this would be the first time that multiple thresholds model approach will be applied to asthma diagnosis in children and adolescents.

Potential limitations emerge from the underreporting of thresholds in existing studies or lack of data for some subgroup analyses, which could narrow the scope of our meta-analysis. This analysis would benefit from obtaining individual patient data from selected studies, though retrieving this data is an extensive and complex process requiring dedicated ethical and digital solutions. As well, we acknowledge that the clinical overlap between atopic conditions, anti-allergic medication, nasal polyposis, and asthma represents significant confounding factors for FeNO and BEC. These challenges will be addressed by our choice of subgroup and sensitivity analysis.

In conclusion, by synthesising existing evidence on the diagnostic accuracy of FeNO alone or combined with BEC in asthma, we intend to offer valuable insights into optimising diagnostic pathways and potentially reducing reliance on invasive tests. This protocol will clarify the potential diagnostic ability of FeNO with or without BEC when traditional diagnostic methods are within normal values.

## Data Availability

All data produced in the present study are available upon reasonable request to the authors

## Registration

In accordance with reporting guidelines, the systematic review protocol was registered with the International Prospective Register of Systematic Reviews (PROSPERO) on 18/12/2023 and was last updated on 31/05/2024 (registration number CRD42023489738)

## SUPPLEMENTARY MATERIAL S1

### Draft search strategy - Ovid MEDLINE(R) ALL <1946 to June 2024>

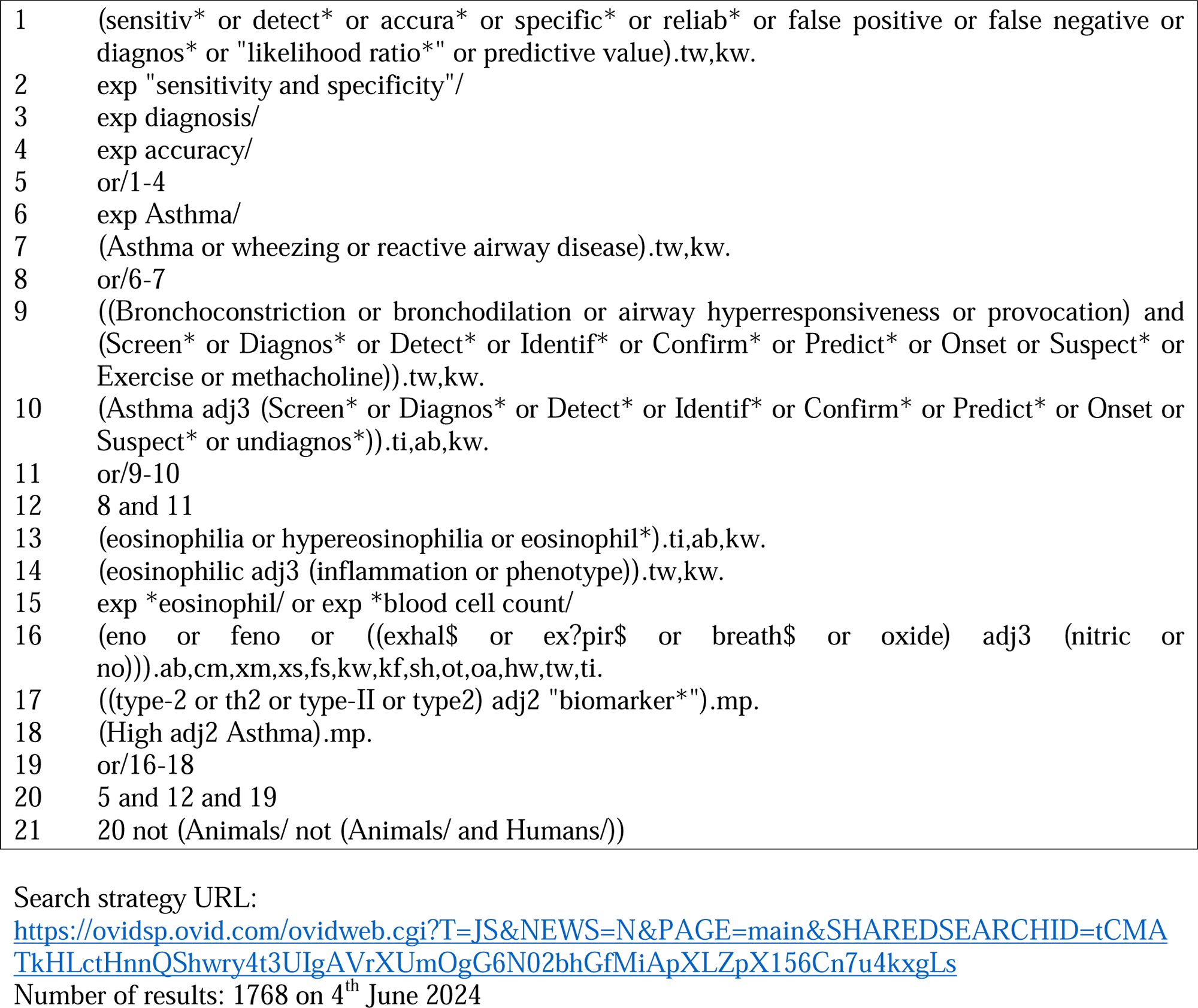

